# Specific Thoracic Pain: Red flag screening pathway for Physiotherapists. A Systematic Review

**DOI:** 10.1101/2021.02.23.21250954

**Authors:** Monica Erbesato, Firas Mourad, Fabrizio Brindisino, Mattia Salomon, Giacomo Rossettini, Michele Margelli, Valerio Barbari, Lorenzo Storari, Federico Andreoletti, Jacopo Disarò, Filippo Maselli

## Abstract

The lifetime prevalence of isolated pain in the thoracic spine is relatively low, approximately 13-17%, compared to neck and low back pain, 40% and 57% respectively. However a patient with thoracic or chest pain is more likely to mask a serious pathology, such as spinal fracture, spinal tumor or metastasis, myocardial ischemia, pneumonia, etc.. A physical therapist is a primary healthcare professional and it is his responsibility to screen and identify serious pathologies outside the scope of practice. That is, differential diagnosis may help in excluding medical pain sources and, most importantly, recognize emergencys scenarios. An in-depth subjective and objective examination are the two pillars for the clinical evaluation that may help clinicians to determine if the pain is of musculoskeletal origin or not. In the literature, there is a lack of knowledge about the red flags (i.e. warning symptoms and signs) that can be useful for the screening for referral, not medical diagnosis, in the thoracic and chest wall pain. The aim of our systematic review is to investigate which findings form the subjective (symptoms and red flags) and objective examination (signs and tests) are valid tools for the differential diagnosis for thoracic pain or chest pain suspected to be caused by a serious pathology.

## Background

According to the current definition, thoracic spinal pain is pain reported anywhere within the region bounded superiorly by the first thoracic spinous process, inferiorly by the last thoracic spinous process, and by the most lateral margins of the erector spinae muscles (Harold Merskey, 1994). Chest pain is defined as pain reported anywhere in the anterior chest wall (region bounded superiorly by the thoracic outlet, inferiorly by the diaphragmatic margin, and laterally by the mid-axillary line) (Mette J Stochkendahl, 2008). With the term ‘dysfunction’ the authors recognize impairment in the musculoskeletal system of Thoracic Spine which may affect its integrity during functional movement; a synergy of motion occurring across different joints. The lifetime prevalence of isolated pain in the thoracic spine is relatively low, approximately 13-17%, compared to neck and low back pain, 40% and 57% respectively (Andrew M Briggs, 2009). However thoracic and chest wall pain is more likely to be caused by serious underlying pathology than neck or low back pain. Therefore, a patient complaining of thoracic or chest pain is more likely to mask a serious pathology, such as spinal fracture, spinal tumor or metastasis, myocardial ischemia, pneumonia, etc.. As physical therapist is primary healthcare professional, it is his responsibility to screen and identify serious pathologies outside the scope of practice. That is, differential diagnosis may help in excluding medical pain sources and, most importantly, recognize emergency’s scenarios.An in-depth subjective and objective examination are the two pillars for the clinical evaluation that may help clinicians to determine if the pain is of musculoskeletal origin or not. In the literature, there is a lack of knowledge about the red flags (i.e. warning symptoms and signs) that can be useful for the screening for referral—not medical diagnosis--in the thoracic and chest wall pain. The aim of our systematic review is to investigate which findings form the subjective (symptoms and red flags) and objective examination (signs and tests) are valid tools for the differential diagnosis for thoracic pain or chest pain suspected to be caused by a serious pathology. We will include all the findings used by all kind of Healthcare Professional, different healthcare setting that were compared compared to the gold standard, (i.e. diagnostic imaging, laboratory testing and biopsy examination) possessing information of their diagnostic accuracy and their psychometric properties.

## METHODS

This systematic review will be conduct in accordance with the Preferred Reporting Items for Systematic Reviews and Meta Analyses (PRISMA) statement (htt). This review will be registered on Prospero Database. The following databases will be searched: Medline (PubMed), Embase, Cochrane Library, DITA, Google Scholar, CINAHl. The search will be limited to English language publications only. No study design or language limits will be imposed on the search. In addition, a manual search will be performed on the reference lists of included articles and other grey literature sources (eg. Google Scholar). References list of identified articles and reviews will be also checked for any relevancy

### Inclusion and exclusion criteria

#### Participants/population

Partecipants will be adults (≥ 18years), suffering of thoracic pain and/or chest pain and/or and/or Upper back pain and/or thoracic dysfunctions. Studies will be considered eligible only if includes detailed information form the subjective (symptoms and red flags) and objective examination (signs and tests), included their psychometric properties collected during both primary or secondary care settings. Patients with a previous diagnosis of serious pathology will be excluded.

#### Intervention, exposure

This review focuses on al, the relevant findings from the subjective (symptoms and red flags), objective examination (signs and tests), and diagnostic/screening tools for the triage of serious spinal pain performed by primary and secondary of different healthcare professionals, such as physiotherapist, cardiologist, pneumologist, rheumatologist and oncologist, etc. All tests or tools without psychometric properties or gold standard comparison will be excluded.

#### Comparator/control

The comparison will be diagnostic imaging and biopsy examination. Studies will be included if the presence or absence of chest pain and/or chest pain and/or chest dysfunction and/or low back pain of different origins was confirmed by a procedure considered the reference standard for the diagnosis of a given pathological condition. By reference standard all diagnostic images were considered - e.g. radiography, computerized tomography (CT) scan, magnetic resonance imaging (MRI), bone scintigraphy, ultrasound - (laboratory tests, surgical findings or any other investigation that objectively certifies the presence or not of pathology) (we will evaluate whether to put these).

#### Studies design selection

We will include only primary studies: diagnostic accuracy studies and longitudinal observational studies, e.g. cohort and case-control studies. Instead case report and case series such as systematic review will be excluded.

#### Main outcomes to investigate

The main outcome is to describe the psychometric properties measured by the comparison of a gold standard of the most relevant findings from the subjective (symptoms and red flags), objective examination (signs and tests) able to be implemented in routinely clinical practice with the goal of triage of patients with thoracic or chest pain of specific origin. Adjunctive outcomes are to relate to the primary outcome the diagnosis of the serious pathology, to identify the healthcare professional performing the screening or diagnostic tool and the setting in which those tests are performed.

We will include different type of effect measure:

- Odds ratio to determine association of specific screening or diagnostic tool with the diagnosis of a serious pathology
- Sensitivity, Specificity, positive likelihood ratio (LR+), negative likelihood ratio (LR-), positive and negative predictive value (PPV and NPV) in detecting serious pathology by screening or diagnostic tool used by healthcare professional.

#### Search strategy and Data extraction

To answer our clinical question and to investigate the scientific literature we will built the searching strategy using both medical subject headings (MeSH) and free terms--entry terms included--related to thoracic and chest pain (for Population), diagnostic tool, screening tool and diagnostic test (for Examination/Intervention) and all kind of diagnostic imaging (for Comparison). Then we will adapt the terms and the syntax to the other electronic database, that are previously mentioned. Two reviewers will screen independently titles and abstracts found in the databases to determine the eligibility of the studies. After the selection and the inclusion of the studies two reviewers will extract and organize in Excel sheet and Word document for further analysis and synthesis of the data. A second independent reviewer will check and verify the extracted data. If consensus is not reached, a third independent reviewer will help to resolve any discrepancy. To evaluate the internal validity and methodological rigor of the included studies we will use JBI tool for cross sectional study, case control and cohort studies; in addition the QUADAS 2 for diagnostic study will be used. We will also analyze the results splitting all the data into subgroups in order to create comparisons between them. The data will be divided into subgroups according to the healthcare professional that performed the evaluation.

#### Risk of bias (quality) assessment

To evaluate the internal validity and methodological rigor of the included studies we will use JBI tool for cross sectional study, case control and cohort studies; in addition the QUADAS 2 for diagnostic study will be used.

#### Strategy for data synthesis

After the appraisal process, studies with adequate internal validity and methodological rigor will be considered admissible and will be included in the analysis. Evidence tables and summary of findings tables will be produced.

#### Analysis of subgroups or subsets

_We will also analyze the results splitting all the data into subgroups in order to create comparisons between them. The data will be divided into subgroups according to the healthcare professional that performed the evaluation.

## RELEVANCE AND DISSEMINATION

We believe that results in this systematic review will increase clinicians knowledge about symptoms and signs that may mimic a serious pathology. We hope that the results of this systematic review will propose clinicians the most reliable screening test to detect a serious pathology.

## Data Availability

All data results will be published.

